# Evaluation of submandibular gland biopsies with RT-QuIC in Parkinson’s disease under investigation

**DOI:** 10.1101/2025.10.22.25337489

**Authors:** Wiebke M. Jürgens-Wemheuer, Daniel Martens, Jörg Spiegel, Leja Nessis, Phillip Kulas, Lukas Pillong, Florian Rosar, Konstantin Redl, Annika Lechler, Arne Wrede, Klaus Faßbender, Walter J. Schulz-Schaeffer

## Abstract

**Background:** The definite diagnosis of Parkinson’s disease (PD) is usually made by the detection of α-synuclein aggregates in the brain *post mortem* with the rare exception of some genetic forms. Traces of α-synuclein aggregates in extracerebral tissue biopsies may serve as an appropriate biomarker to confirm a clinical diagnosis *in vivo*.

**Objectives:** We set out to determine whether the detection of α-synuclein aggregates in submandibular gland biopsies is an effective means for diagnosing PD patients.

**Methods:** We examined submandibular gland biopsies from 25 patients with PD under investigation and 25 age-matched controls to detect α-synuclein aggregates using real-time quaking induced conversion (RT-QuIC) as a seed amplification-assay and immunohistochemical α-synuclein aggregate detection and paraffin-embedded tissue blot (PET-blot) as confirmatory methods.

**Results:** Our RT-QuIC assay detected α-synuclein aggregates in submandibular gland biopsies with a sensitivity of 81,1%, which increased to 90% after a clinical follow-up, and a specificity of 100%. The PET-blot with mAb5C12 confirmed 50% of the RT-QuIC positives and offered a sensitivity of 45,8% (50% after clinical follow-up) and a specificity of 100%. Immunohistochemical detection using the same antibody confirmed 28% of the RT-QuIC positives, but found two of the controls to be positive and therefore provided a sensitivity of 26,1% (28,6% after clinical follow-up) and a specificity of 92%.

**Conclusions:** The RT-QuIC Assay demonstrated comparable sensitivity to the clinical diagnosis (when neuropathologic examination represents the gold standard) and exhibited a similar level of specificity as the PET-blot.

## Introduction

Parkinson’s disease (PD) is the second most common neurodegenerative disease of the elderly population worldwide.^1^ Nevertheless, the diagnosis remains challenging, especially in patients to be diagnosed at early stages.^2^ While an early and accurate diagnosis is undoubtedly favorable for optimal patient management and therapy trials, it is not uncommon for a delay of 12-29 months to occur between the contact with a doctor due to symptoms and the clinical diagnosis by an expert for numerous reasons.^3-7^ Neuropathologic investigations of deceased PD patients show a clinical misdiagnosis for approximately 20%.^8^ For patients at an early disease stage, the accuracy of the diagnosis is even lower at 58%.^9^ Improved diagnostic criteria, follow-up examinations, and the aid of advanced neuroimaging increase the certainty of the diagnosis. However, it is the evidence of α-synuclein aggregates that will confirm the clinical diagnosis of an iatrogenic PD patient *post mortem*. Synaptic α-synuclein aggregates are assumed to be associated with the pathophysiology of PD^10, 11^ and α-synuclein aggregates represent the major component of Lewy bodies, which are the neuropathologic hallmark of PD. Some genetic forms may not comprise Lewy bodies in the brain.^12^ Synaptic α-synuclein aggregates, Lewy bodies and Lewy neurites are inevitable present in the central nervous system of idiopathic PD patients and can also be found to a lesser degree in the periphery.^13^ This favors α-synuclein aggregates as a lead candidate PD biomarker. Extracerebral suitable tissue biopsies could be a valuable tool to confirm the diagnosis *in vivo*.

There has been considerable progress made in the last decade to detect minute amounts of protein aggregates. Techniques originally developed for prion research, such as the paraffin-embedded tissue-blot (PET-blot) and seed amplification assays (SAAs)^14^, including real-time quaking induced conversion (RT-QuIC) have been successfully adapted for the detection of aggregated α-synuclein. The PET-blot conveys the specific detection of α-synuclein aggregates in tissue samples, due to the extensive proteinase K (PK) digestion overnight.^10, 11^ SAAs assume that tiny aggregated pathological protein seeds are sufficient to corrupt added protein monomers into the pathological form, thereby creating an exponential increase of protein aggregates. Over the past eight years, different α-synuclein SAAs have been successfully applied to detect α-synuclein aggregates in cerebrospinal fluid (CSF), blood serum, autopsy and biopsy tissues.^15-19^

Conventional immunohistochemical detection in biopsies and autopsy tissue has definitively shown that an α-synuclein pathology can be found in the submandibular gland of PD patients.^20, 21^ Studies of biopsies of the submandibular glands of patients with PD and of control subjects revealed sensitivity rates that varied from 56.1% to 75%, with specificity rates ranging from 78% to 92.9%.^20-22^ Currently, there is no research available on α-synuclein SAAs or PET-blot studies on actual submandibular gland biopsies. Seed amplification assay studies have so far only been performed on autopsy tissue from the submandibular gland, albeit with a promising sensitivity of 100% and a specificity of 94%.^23^ In the living patient, α-synuclein seed amplification assays have been mainly focused on cerebrospinal fluid (CSF) and skin samples^24^, producing promising results and an FDA-approved official α-synuclein test for CSF in the USA.^14^ However, tissue biopsies in general offer the possibility to evaluate a sample with several techniques.^18^ We sought to determine the suitability of the submandibular gland to serve as a target for the detection of α-synuclein aggregates using the RT-QuIC, an immunohistochemical detection and the PET-blot method.

## Material and Methods

### Patient biopsies of the submandibular gland

We enrolled 25 patients with clinical Parkinson’s disease under investigation at the clinic for neurology and recruited age-matched controls (n = 25) from the clinic for otorhinolaryngology (Saarland University Medical Center), which had surgical procedures on the submandibular gland that were unrelated to the study. Each participant provided written consent for the use of data as well as the biopsy or use of excess tissue, respectively. All patients and their age-matched controls are listed in Table 1. The project was approved by the ethics committee of the Saarland Medical Association (No. 234/19).

**Table 1:** PD patients with their age-matched controls. Patients (P = {P_i_ : i = 1, …, n}) and controls (K ={Kj : j = 1, …, m}) were age-matched using a bipartite graph G = (V= P ∪ K, E), where patients P and controls K form the disjoint set of vertices V and E is the set of all edges between P and K. The weight of an edge is defined as the age difference d_ij_ =|age(P_i_) − age(K_j_)|. The minimum-weight maximal matching algorithm computes a maximal subset E’ of E where the sum of age differences of all edges in E’ is minimal. To prevent outliers, we used the weights w_ij_ = d_ij_^s^ = |age(P_i_) − age(K_j_)|^s^ with s=2. To minimize the risk of identification the actual age of each person has been replaced with an age range.

| PD patient | age (years) | age-matched control | age (years) | age difference (years) |
| --- | --- | --- | --- | --- |
| #P 1 | 71-75 | #C 22 | 71-75 | 3 |
| #P 2 | 61-65 | #C 1 | 56-60 | 5 |
| #P 3 | 71-75 | #C 10 | 71-75 | 3 |
| #P 4 | 66-70 | #C 16 | 61-65 | 5 |
| #P 5 | 66-70 | #C 8 | 66-70 | 1 |
| #P 6 | 46-50 | #C 5 | 46-50 | 2 |
| #P 7 | 56-60 | #C 9 | 51-55 | 2 |
| #P 8 | 66-70 | #C 3 | 66-70 | 2 |
| #P 9 | 66-70 | #C 4 | 61-65 | 5 |
| #P 10 | 76-80 | #C 18 | 71-75 | 6 |
| #P 11 | 76-80 | #C 17 | 71-75 | 5 |
| #P 12 | 66-70 | #C 20 | 61-65 | 2 |
| #P 13 | 66-70 | #C 2 | 56-60 | 7 |
| #P 14 | 61-65 | #C 11 | 56-60 | 3 |
| #P 15 | 71-75 | #C 12 | 66-70 | 3 |
| #P 16 | 81-85 | #C 14 | 81-85 | 3 |
| #P 17 | 71-75 | #C 15 | 66-70 | 2 |
| #P 18 | 51-55 | #C 24 | 51-55 | 1 |
| #P 19 | 66-70 | #C 6 | 61-65 | 3 |
| #P 20 | 66-70 | #C 23 | 66-70 | 2 |
| #P 21 | 51-55 | #C 13 | 51-55 | 0 |
| #P 22 | 51-55 | #C 25 | 51-55 | 0 |
| #P 23 | 76-80 | #C 7 | 76-80 | 4 |
| #P 24 | 71-75 | #C 19 | 66-70 | 3 |
| #P 25 | 76-80 | #C 21 | 71-75 | 5 |

Clinical examination of all patients with suspected Parkinson’s disease was carried out by a neurologist specialized for movement disorders. The diagnosis was made in accordance with the guidelines of the German Society for Neurology^24^ and the recommendations of the International Parkinson and Movement Disorder Society (MDS) diagnostic criteria for diagnosing PD^25^. Imaging diagnostics were performed on 16 patients at an early stage of the disease; primarily utilizing techniques such as dopamine transporter targeted single photon emission computed tomography (DaT-SPECT) using ^123^I-FP-CIT and ^18^F-fluorodeoxyglucose positron emission tomography (FDG-PET) (Table 2).

**Table 2:** Results for RT-QuIC, PET-blot and immunohistochemical detection (IHC) of all patients with Parkinson’s disease under investigation and controls in this study, including the time span between diagnosis and biopsy. To minimize the risk of identification the actual age of each person has been replaced with an age range.

| No. | sex | age | patient/<br>control | clinical disease<br>years | PD type | DaT-SPECT | RT-QuIC | PET-blot | IHC |
| --- | --- | --- | --- | --- | --- | --- | --- | --- | --- |
| #P 1 | f | 71-75 | patient | 28 | hypokinetic-rigid | none | positive | negative | negative |
| #P 2 | f | 61-65 | patient | 21 | equivalent type, plus camptocormia | none | positive | negative | positive |
| #P 3 | f | 71-75 | patient | 20 | equivalent type | none | n.a. | positive | negative |
| #P 4 | m | 66-70 | patient | 20 | hypokinetic-rigid | none | positive | positive | negative |
| #P 5 | f | 66-70 | patient | 19 | hypokinetic-rigid | none | n.a. | positive | positive |
| #P 6 | m | 46-50 | patient | 18 | equivalent type | positive | positive | negative | negative |
| #P 7 | m | 56-60 | patient | 15 | hypokinetic-rigid | none | positive | positive | negative |
| #P 8 | m | 66-70 | patient | 15 | hypokinetic-rigid | positive | positive | positive | negative |
| #P 9 | m | 66-70 | patient | 15 | tremor dominant | positive | positive | negative | negative |
| #P 10 | m | 76-80 | patient | 14 | hypokinetic-rigid, plus<br>camptocormia | none | positive | positive | positive |
| #P 11 | m | 76-80 | patient | 14 | hypokinetic-rigid | none | positive | positive | positive |
| #P 12 | f | 66-70 | patient | 14 | equivalent type, plus on-off dystonia | none | positive | n.e. | n.e. |
| #P 13 | m | 66-70 | patient | 12 | hypokinetic-rigid | positive | positive | negative | negative |
| #P 14 | m | 61-65 | patient | 12 | hypokinetic-rigid | negative | negative | negative | negative |
| #P 15 | m | 71-75 | patient | 12 | equivalent type | positive | positive | positive | negative |
| #P 16 | f | 81-85 | patient | 12 | hypokinetic-rigid | none | positive | positive | positive |
| #P 17 | f | 71-75 | patient | 10 | tremor dominant | none | negative | negative | negative |
| #P 18 | m | 51-55 | patient | 7 | hypokinetic-rigid | none | positive | negative | negative |
| #P 19 | m | 66-70 | patient | 4 | tremor dominant | positive | negative | negative | negative |
| #P 20 | f | 66-70 | patient | 4 | tremor dominant | positive | positive | positive | negative |
| #P 21 | m | 51-55 | patient | 4 | hypokinetic-rigid | negative | negative | negative | negative |
| #P 22 | m | 51-55 | patient | 3 | hypokinetic-rigid | none | n.a. | negative | negative |
| #P 23 | m | 76-80 | patient | 1 | hypokinetic-rigid | none | positive | positive | n.e. |
| #P 24 | m | 71-75 | patient | 1 | hypokinetic-rigid | positive | positive | negative | positive |
| #P 25 | f | 76-80 | patient | 1 | equivalent type | none | positive | negative | negative |
| #C 1 | f | 56-60 | control |  |  |  | negative | negative | negative |
| #C 2 | m | 56-60 | control |  |  |  | n.a. | negative | negative |
| #C 3 | f | 66-70 | control |  |  |  | n.a. | negative | negative |
| #C 4 | m | 61-65 | control |  |  |  | negative | negative | negative |
| #C 5 | m | 46-50 | control |  |  |  | negative | negative | negative |
| #C 6 | f | 61-65 | control |  |  |  | negative | negative | negative |
| #C 7 | f | 76-80 | control |  |  |  | negative | negative | negative |
| #C 8 | f | 66-70 | control |  |  |  | negative | negative | negative |
| #C 9 | f | 51-55 | control |  |  |  | negative | negative | negative |
| #C 10 | m | 71-75 | control |  |  |  | negative | negative | negative |
| #C 11 | m | 56-60 | control |  |  |  | negative | negative | negative |
| #C 12 | f | 66-70 | control |  |  |  | negative | negative | negative |
| #C 13 | m | 51-55 | control |  |  |  | n.a. | negative | positive |
| #C 14 | m | 81-85 | control |  |  |  | n.a. | negative | positive |
| #C 15 | f | 66-70 | control |  |  |  | negative | negative | negative |
| #C 16 | f | 61-65 | control |  |  |  | negative | negative | negative |
| #C 17 | m | 71-75 | control |  |  |  | n.a. | negative | negative |
| #C 18 | f | 71-75 | control |  |  |  | negative | negative | negative |
| #C 19 | f | 66-70 | control |  |  |  | n.a. | negative | negative |
| #C 20 | f | 61-65 | control |  |  |  | negative | negative | negative |
| #C 21 | f | 71-75 | control |  |  |  | negative | negative | negative |
| #C 22 | f | 71-75 | control |  |  |  | negative | negative | negative |
| #C 23 | m | 66-70 | control |  |  |  | negative | negative | negative |
| #C 24 | m | 51-55 | control |  |  |  | negative | negative | negative |
| #C 25 | m | 51-55 | control |  |  |  | negative | negative | negative |

n.a. = not available; n.e. = not evaluable

Patients received local anesthesia before a small stab incision was made and the 14G needle (Max-Core®, BD, Switzerland) was used to gain the submandibular tissue in caudo-cranial direction under sonographic control. The skin lesion was closed and followed by another sonographic control to detect unexpected bleeding.

### Autopsy tissues

We used already obtained autopsy material from a larger study on Parkinson’s disease, which has been approved by the ethics committee of the Medical Association of Hessen, Germany (Nr. FF89/2008). Material included frozen tissue and paraffin-embedded tissue sections from brain and submandibular gland samples (Suppl. table 1) and was used to establish methods and provide controls.

### Tissue preparation

Biopsies and excess tissue from the otorhinolaryngology unit were divided for cryopreservation at −80 °C and fixation with 4% buffered paraformaldehyde for 48 hours.

Formalin-fixed tissue was embedded in paraffin using a 16-hour protocol for small tissue samples. We cut 1-3 µm paraffin sections and placed them on glass slides (SuperFrost Plus, Menzel GmbH & Co KG, Germany) or on nitrocellulose membranes, pore size 0.45µm (16201150, BioRad, Hercules, USA). Sections on glass slides were routinely stained with hematoxylin/eosin (H&E), and elastica van Gieson (EVG) for morphological examination.

### Real-time quaking induced conversion assay (RT-QuIC)

We followed the streamlined protocol of Bargar et al.^26^ with few modifications from Pinder et al.^27^, using aliquoted 10% homogenates of brain samples and of the submandibular gland. Nunc black 96-well plates with optical flat bottom (Thermo-Fisher, Waltham, USA) were preloaded with six 0.8mm silica beads per well (OPS-Diagnostics, Lebanon, USA) plus 98µl of the reaction mixture containing 170mM NaCl, 400mM phosphate buffer (pH 8, the stock solution consisting of 46.6ml of 1M Na_2_HPO_4_ and 3.4ml of 1M NaH_2_PO_4_), 20µM ThioflavinT (T-3516, Sigma, St. Louis, USA) and 0.1mg/ml recombinant α-synuclein monomers (S1001-2, batches #070921AS and #51722AS, r-peptide, Watkinsville, USA). Before use, the latter was filtered through Amicon 100 kDa filters (Merck Millipore, Tullagreen, Ireland) by centrifugation (10 minutes, 14.000*g*). Tissue sample dilutions were prepared in phosphate buffered saline (AM9625, Thermo-Fisher Scientific, Vilnius, Lithuania) supplemented with Gibco 1×N2 supplement (Thermo-Fisher, Waltham, USA), where the 10% homogenate represented the 10 ^-1^ dilution. Sample dilutions were added in quadruplicates of 2µl and the plate was sealed (Nunc clear sealing film, Thermo-Fisher, Waltham, USA). Plates were incubated at 42°C in a BMG Labtech FLUOstar Omega (RRID:SCR_025024) with cycles of one-minute shaking (600rpm, double orbital) and one-minute rest cycles for 48 to 72 hours. ThT-fluorescence (450nm excitation and 480nm emission; bottom read) was recorded every two minutes and the reader was set up with an optic gain coefficient of ∼1600. Our baseline of ThT-fluorescence was around 30,000 relative fluorescence units (rfu) with the maximal fluorescence response of the plate reader fixed to 260,000rfu. A sample was considered positive when three or four of the quadruplicates showed a positive reaction, defined as enhanced ThT fluorescence above the threshold. The threshold was calculated as the average background fluorescence plus five standard deviations, resulting in a threshold value of ∼ 52.000 rfu. The lag phase of RT-QuIC reactions was defined as the time (hours) to cross this threshold (Figure 1F). A sample with two out of four positives was repeated while one or none positive reaction was considered to be negative.

**Figure 1:**
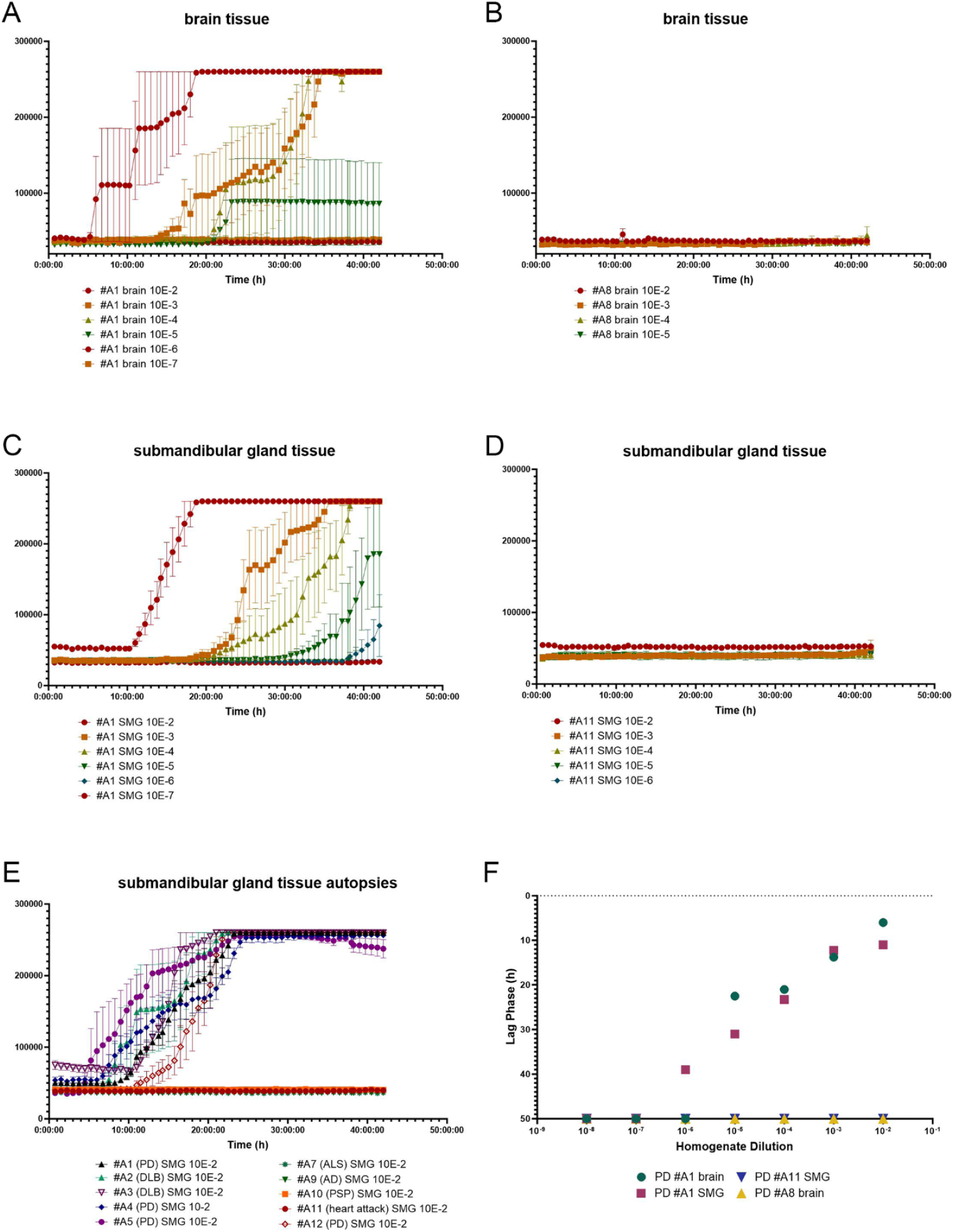
Autopsy samples were used to establish the RT-QuIC protocol for brain and submandibular gland (SMG) tissue as exemplary shown (A-D) with dilutions ranges from 10^−2^ to 10^−7^ and the corresponding lag phases in hours (F). The lowest SMG dilution 10^−2^ was consistently positive for all PD and DLB autopsy cases and negative for control samples (E). Error bars indicate the s.e.m. for the four quadruplicates of each sample

Data was analyzed using MARS Data Analysis Software (RRID:SCR_021015), Microsoft Excel (RRID:SCR_016137), and GraphPad Prism (RRID:SCR_002798).

We established the RT-QuIC using brain samples in dilutions ranging from 10^−2^ to 10^−7^ as exemplified in Figure 1 (A, B). Once the RT-QuIC worked for brain tissue, we moved on to submandibular gland tissue (Figure 1C and D) and generated a homogenate of all available biopsy tissue for each patient. This approach ensured that the source material remained consistent for repetitions. Frozen aliquots of tissue homogenates and reconstituted protein were only used once. Different tissue samples from the same autopsy gland had always produced the same result in our hands. For submandibular gland autopsy samples, the lowest dilution 10^−2^ reliably produced a positive reaction in all four wells within 13.5 hours (Figure 1E and 2C), provided there were no major fluctuations regarding the room temperature. Therefore, we tested all available submandibular gland biopsies at this dilution (Figure 2A and B). Due to the previously described inhibitory effect of negative brain homogenate on the RT-QuIC^33^, which we could also observe (data not shown), we refrained from calculating supposed sensitivities in a comparison of brain and submandibular tissue. Having finished all experiments, neuropathologists and scientific staff were de-blinded by the neurologists.

**Figure 2:**
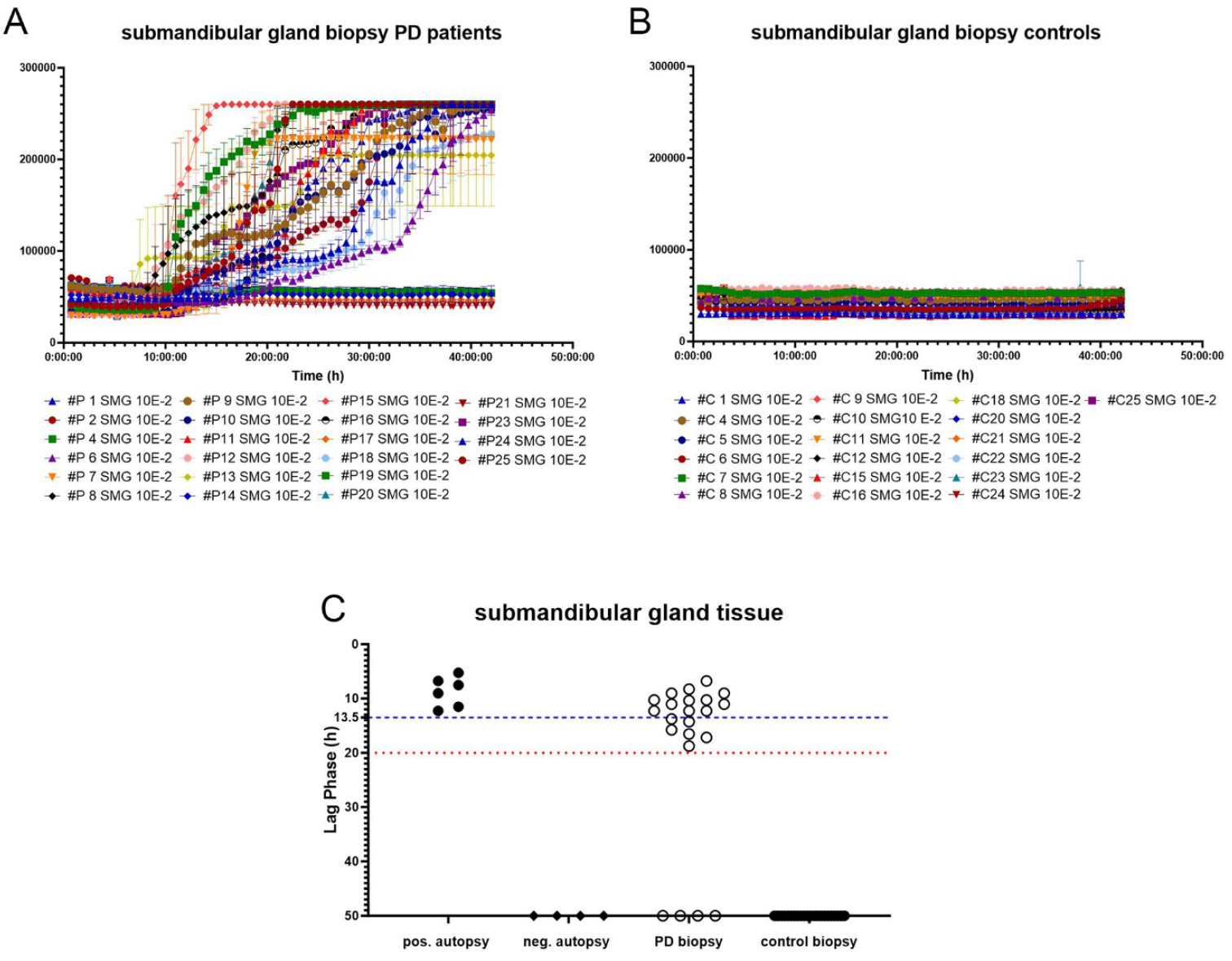
Since the lowest dilution 10^−2^ was consistently positive for all PD and DLB autopsy cases, we tested all biopsy specimens from clinical PD patients (#P) and controls (#C) at 10^−2^ (A-C). Positive biopsies crossed the threshold within 20 hours (red dotted line, C) and positive autopsy samples within 13.5 hours (blue dashed line in C), while controls and negative samples stayed negative for at least 50 hours (C).

### Paraffin-embedded tissue blot (PET-blot)

The PET-blot was carried out as described previously.^10^ Sections on membranes were deparaffinized in a series of xylene, isopropanol (instead of ethanol), washed in PBST [PBS with 1% Tween® 20 (Polysorbate 20, #91272, Roth, Germany)] and dried overnight. We used a proteinase K (PK) concentration of 250µg/ml at 56°C and the pillow technique where the pre-wetted membranes are placed overnight on tissues soaked with PK-buffer [Tris-buffered saline pH 7.8 with Brij® (B4184, Merck KGaA, Germany)] and PK (P6556, Sigma-Aldrich, St. Louis, USA). This procedure ensures that protein aggregates remain and it prevents them from floating off in puddles of buffer. Sections were submitted to denaturation with 4M GdSCN (#0017.2, Carl Roth GmbH, Germany) for 15 minutes and blocking with 0.2% casein (I-Block™, T2015, Applied-Biosystems, Foster City, USA) for 45 minutes. All steps were carried out with gentle agitation at room temperature and membranes were rinsed with TBST in-between. Primary antibody 5C12 against unmodified α-synuclein (epitope: amino acids 109–120, 1.5mg/ml, Prothena Biosciences Limited, Ireland) was applied at a dilution of 1:10.000 in TBST for 90 minutes followed by a secondary alkaline phosphatase-coupled antibody (ab6790, abcam, UK) at 1:2000 for 60 minutes. Membranes were rinsed with NTM buffer (100mM NaCl, 100mM Tris pH 9.5, 50mM MgCl_2_) to adjust the pH in preparation for the formazan reaction with NBT/BCIP substrate, which was conducted under visual control of a brain control section and stopped in PBS. The resulting dark purple chromogen was evaluated under a dissection microscope).

### Immunohistochemical detection

After deparaffinization and rehydration, we treated tissue slides with a peroxidase block containing 1% H_2_O_2_. They were subjected to PK digestion at 50µg/ml for 15 minutes in pre-warmed 37 °C PK-buffer (see PET-blot section). After rinsing (Dako Wash Buffer 10x, #S3006, Santa Clara, USA), unspecific reactions were blocked with 0.2% casein (see above) for 10 minutes. Antibody 5C12 was applied 1:1000 diluted in TBS for 45 minutes at room temperature in a humidity chamber. We rinsed the slides and incubated them for 30 minutes with Dako Real EnVision HRP Rabbit/Mouse (ref #K5007, Santa Clara, USA). The antigen-antibody reaction was visualized using Dako REAL DAB + Chromogen (50X, K5007, Santa Clara, USA) according to the manufacturer’s instructions. Sections were lightly counterstained with hematoxylin, dehydrated and coverslipped.

Images of tissue sections on membrane or glass slides were taken with a U-TV0.5XC-3 microscope (Olympus, Tokyo, Japan). Adjustments to the white balance were made using Adobe Photoshop (RRID:SCR_014199), which was also used to stack the PET-blot images when necessary.

## Results

### Clinical characterization of patients with Parkinson’s disease under investigation

Of the 25 patients with clinical Parkinson’s disease under investigation (mean age 67.3 ± 9.3 years s.d.), sixteen were male (mean age 64.8 ± 10.2 years s.d.) and nine were female (mean age 71.4 ± 6 years s.d.). The age-matched (Table 1) control group (mean age 64.6 ± 8.6 years s.d.) consisted of 12 men (mean age 62.8 ± 10.6 years s.d.) and 13 women (mean age 66.4 ± 6.2 years s.d.). Table 2 provides clinical data and the results of the α-synuclein aggregate detection for each participant. Not all samples were available for all α-synuclein aggregate detection methods. The mean time span between diagnosis and biopsy was 11.8 ± 7.2 years (s.d.).

A clinical follow-up was conducted approximately four years after the biopsy was taken in the patients where we could not find α-synuclein aggregates. Of these four patients, two met the MDS criteria for the diagnosis of PD (#P 17 and #P 19). Both showed disease progression and their symptoms responded to dopaminergic medication. Additionally performed diagnostics including transcranial sonography and FDG-PET further supported the diagnosis of PD. Patient #P 17, however, could be a case of familial PD based on anamnestic evidence in the family history. No genetic testing has been conducted thus far. The two patients #P 14 and #P 21 showed mild symptoms and no disease progression over 16 and eight years, respectively. In accordance with the MDS criteria, a re-evaluation of the diagnosis is recommended in such patients. Here, the clinical examination was extended to a recalculation of the Z-ratio of DaT-SPECT results (by additional semiautomatic software), which led to an exclusion of the diagnosis PD for both. Summarized, the follow-up examination led to a revised diagnosis in patients #P 14 and #P 21.

### Detection of α-synuclein aggregates with the RT-QuIC

Our RT-QuIC protocol definitively showed that all control samples (19/19) tested negative (Figure 2B and C). In 18 of 22 patients with Parkinson’s disease under investigation, the RT-QuIC detected α-synuclein aggregates in their submandibular gland biopsies (Figure 2A and C, Table 2). On average, biopsies that tested positive required 12.2 ± 3.3 hours (s.d.) to exceed the calculated threshold, while autopsy tissue from the submandibular gland that tested positive required an average of 8.7 ± 2.7 hours (Student’s t-test, P = 0.0314), reflecting thereby the more intense accumulation of aggregates in terminally ill and deceased patients. Positive biopsy samples crossed the threshold within the first 20 hours (dotted red line in Figure 2C). The individual lag phases for each submandibular sample are displayed in Figure 2C.

The assay’s sensitivity, specificity, and accuracy in detecting α-synuclein aggregates in submandibular biopsy tissue were determined to be 81.8%, 100%, and 90.2%, respectively. After reviewing the follow-up data from approximately four years after the biopsy, it was observed that two of the four RT-QuIC negative patients no longer met the criteria for Parkinson’s disease as previously defined. This resulted in an increase in the sensitivity and accuracy of the RT-QuIC, reaching 90% and 95.1%, respectively. The specificity remained at 100%.

### Comparison of RT-QuIC to morphological detection methods of α-synuclein pathology

For the submandibular gland biopsies, two PET-blot sections from each participant were viewed, and they were considered positive when longitudinal nerve fibers showed distinctive staining (Figure 3 C and D). A comparative analysis of RT-QuIC and PET-blot revealed that 50% of patients who tested positive via RT-QuIC also exhibited positive results with the PET-blot (9/18). In a manner analogous to the RT-QuIC none of the examined control cases (25/25) exhibited PK-resistant α-synuclein aggregates with the PET blot (specificity 100%), while 45.8% (11/24) of the patients with Parkinson’s disease under investigation displayed the presence of few, albeit distinct, thread-like and punctate α-synuclein aggregates (see Figure 3D). Thus, the accuracy of the PET-blot was 73.5%. The revised diagnosis of two patients based on the follow-up examinations resulted in an increase in the sensitivity and accuracy of the PET-blot to 50% and 77.6%, respectively. There was a remarkable quantitative difference in α-synuclein deposition between cortex and submandibular gland in autopsy tissue (Figure 4A and C).

**Figure 3:**
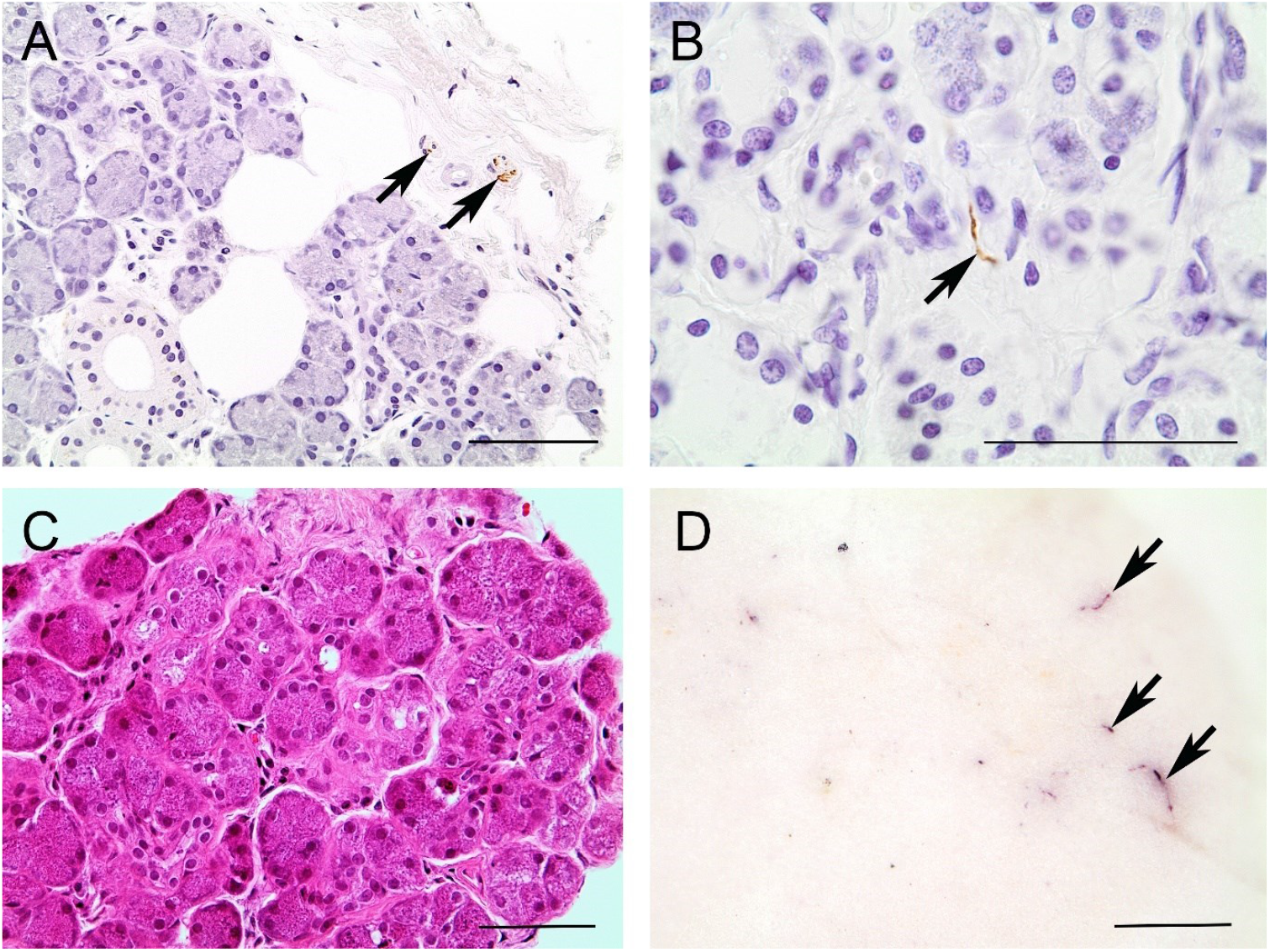
In biopsies of the submandibular gland discrete α-synuclein aggregates may be found in small nerves (A) or between the acini (B) using immunohistochemical detection (mAb5C12 1:1000; bars = 120µm). The PET-blot (D) with the corresponding H&E (C) section reveals distinctive thread-like (D, black arrows) and punctate staining resembling α-synuclein aggregates between the acini (PET-blots mAb5C12 1:10,000; bars = 100µm

**Figure 4:**
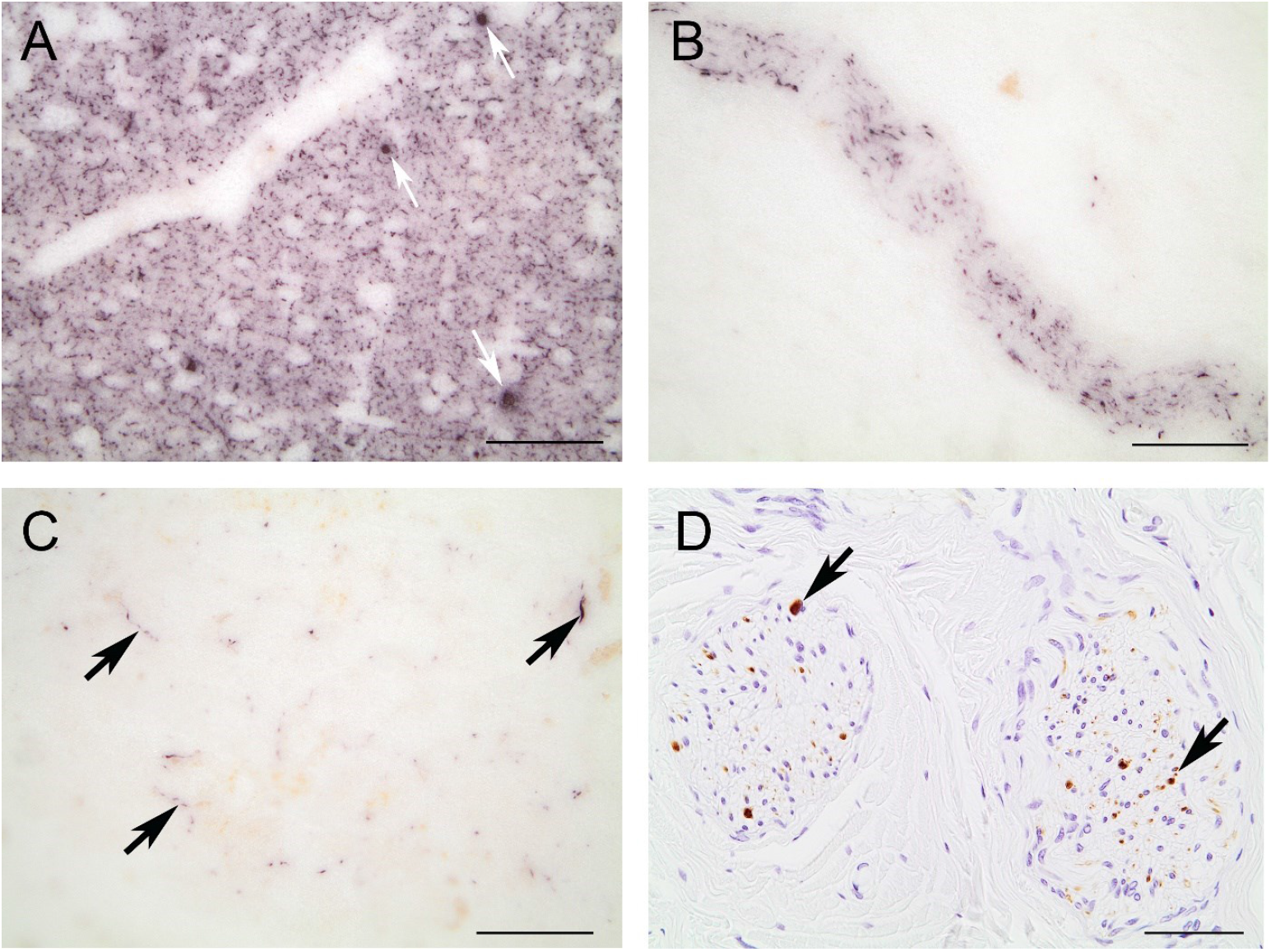
With the PET-blot abundant proteinase K-resistant synaptic α-synuclein aggregates and Lewy bodies (white arrows) are visible in the grey matter of the brain (A), while distinct α-synuclein aggregates can be seen confined to nerves (B) and single nerve fibers between the acini (C, black arrows) in submandibular gland tissue from **autopsies** [PET-blots mAb5C12 1:10,000; bars = 250μm (A, B, C)]. These patterns can be verified by immunohistochemical detection, where distinct α-synuclein aggregates are visible (D, black arrows) in nerves (mAb5C12 1:1000; bar = 120μm).

For immunohistochemical detection of α-synuclein aggregates, we examined one to two biopsy sections that had been stained and considered them positive when distinct staining in nerve fibers (see Figure 3A) or between acini (see Figure 3B) was detectable as previously verified in the autopsy tissue (Figure 4D). Utilizing this method, we identified α-synuclein aggregates in 28% (5/18) of the RT-positive submandibular gland biopsies. In two controls, positive staining was identified where the PET-blot was negative and frozen tissue for RT-QuIC was not available (Table 2). The sensitivity, specificity, and accuracy were therefore calculated to be 26.1% (6/23), 92% (23/25), and 60.4%, respectively, at the time of the biopsy. The revised diagnosis of two patients four years later led to a sensitivity of 28.6% (6/21), a specificity of 92.6% (25/27), and an overall accuracy of 64.6%.

## Discussion

In our diagnostic approach to detect α-synuclein aggregates in submandibular gland biopsies of 25 patients with Parkinson’s disease under investigation and their age-matched controls, we compared RT-QuIC, PET-blot and the immunohistochemical detection. The objective of this study was twofold: first, to ascertain whether the submandibular gland could serve as a suitable target organ to support the clinical diagnosis of PD using α-synuclein aggregates as a biomarker; and second, to validate the applied methods for their sensitivity and specificity in the target tissue.

It was thought that the submandibular gland might be a promising candidate to detect peripheral α-synuclein aggregates, based on the findings of an extensive autopsy study employing immunohistochemical detection of phosphorylated α-synuclein that was conducted in 2010.^13^ Further investigation in PD patients using Meta-Iodo-Benzyl-Guanidine (MIBG) scintigraphy to visualize the sympathetic innervation of organs showed a reduced MIBG accumulation in salivary glands^28^, which lends support to the idea of the submandibular gland being an organ timely involved into pathogenic accumulation of α-synuclein aggregates. In the present study, we observed that RT-QuIC of submandibular gland biopsies demonstrated a sensitivity of 81.8% and a specificity of 100% resulting in an overall accuracy of 90.2%. This is similar to the sensitivity for clinical PD diagnosis when compared to neuropathologic diagnosis.^29^ Studies have confirmed the validation of the MDS criteria with a sensitivity of 94% and a specificity of 88%^25^, which also reflect the degree of uncertainty, particularly in patients in the early stages of the disease with mild motoric symptoms. The diagnostic value of the RT-QuIC even increased to a sensitivity of 90% after the follow-up examinations of patients with uncertain status (negative for α-synuclein aggregates) four years after the biopsy, which led to a revised diagnosis in two cases. While this is reassuring for the reliability of the method, it also corroborates the established finding that the accuracy of clinical PD diagnosis improves over time.^2^ For the patient cohort in this study the submandibular gland seems to be an organ sufficiently affected by α-synuclein pathology for the sensitive detection with the RT-QuIC. According to our experiments, the presence of α-synuclein aggregates would result in a positive RT-QuIC within the first 20 to 24 hours. Yet, the exact point in the pathogenesis of PD when α-synuclein aggregates are detectable in the submandibular gland remains to be determined. Body-first and brain-first type as postulated by Borghammer and colleagues may additionally influence the utility of the submandibular gland as a diagnostic tissue, but to the best of our knowledge, the salivary glands were not examined during their comprehensive investigation of iLBD cases in 2022.^30^

To date, the focus of α-synuclein seeding amplification assays for the *in vivo* approach has been on skin tissue^18, 31^ or liquid biopsies, particularly cerebrospinal fluid^15, 16, 32, 33^, and submandibular gland tissue has only been examined in autopsy cases^23^. The SyNeurGe classification^24^, as established in 2024, considers α-synuclein seeding amplification assays of cerebrospinal fluid and skin biopsies. For the submandibular gland and blood testing, there is currently not enough evidence. For clinically diagnosed PD patients, our RT-QuIC assay could offer confirmation of the α-synuclein status via submandibular gland biopsy, which might therefore become a valid option to be included into the SyNeurGe classification. Further validated submandibular gland biopsies, as intended for the material of the S4 study^21^, may confirm this. The “neuronal α-synuclein disease integrated staging system” (NSD-ISS) proposed by the MJFox foundation, however, considers only the Amprion Test SYNTap, now called SAAmplify-αSYN Test, of the cerebrospinal fluid as evidence for α-synuclein aggregates in vivo.^34^ While the early α-synuclein SAAs employed for the cerebrospinal fluid of living patients took five to 12 days^15, 33^, later protocols were faster^16, 26^, like the present one for submandibular gland analysis. Their sensitivity and specificity ranged from 88.5% to 95% and 96.9 to 100%, respectively. Specifically mentioned, the Amprion Test SYNTap® advertised a sensitivity of 78.7% to 87% and a specificity of 89.5% to 97% depending on whether the clinical PD cohort had been further diagnosed with DaT-SPECT or not [https://ampriondx.com/parkinsons-disease/- *access January 2024*]. In relation to this, the RT-QuIC assay for the submandibular gland with a sensitivity of 90% and specificity of 100% seems to be a valid option, even though a larger cohort would be desirable.

A tissue biopsy facilitates further examination through additional methods. When offering a biopsy to patients with PD, as with any surgical procedure, there are factors to consider, which certainly include the risk of tissue removal and the simplicity of the laboratory procedure. The use of skin biopsies has demonstrated encouraging results *in vivo* and the surgical procedure appears to be less invasive than a biopsy of the submandibular gland. However, typically, even from a single site, such as the cervical neck, at least two biopsies are obtained^18, 31^ to ensure the presence of sufficient nerves, while we could rely on one biopsy that allows biochemical and morphological detection methods. In our study, none of the patients who underwent a biopsy of the submandibular gland suffered from complications afterwards. By morphological detection, the PET blot method was only able to confirm 50% of the RT-QuIC positive biopsies and the immunohistochemical detection of α-synuclein aggregates in the RT-QuIC positives with 28% was indeed very low. In the end, the fact that we used extremely thin and very few tissue sections, thereby examining only a fraction of the paraffin-embedded-tissue, can best explain this. The presence of a limited number of stained nerve fibers may be lost in subsequent sections, which also explains why our immunohistochemistry results and PET-blot results do not always agree for the RT-QuIC positives. By immunohistochemical detection, we found two of the controls to harbor convincingly positive signals. Since the applied PK-digestion was mild when compared to the PET-blot, there might have been residues from physiological α-synuclein. Alternatively, artefacts are known to mimic immunohistochemical DAB deposits.

### Limitations

The study group for our experiments included only 50 participants and we are aware of the fact that not many PD patients were biopsied within the first five years after diagnosis. Therefore, this study is not representative for very early Parkinson’s disease. The setting corresponds more to a verification process once the clinical symptoms recommend the patient to see a neurologist for suspected Parkinson’s disease.

## Data Availability

The datasets generated and analyzed during the current study are included in this published article and its supplementary information files. Additional information and material are available from the corresponding author on reasonable request.

## Code availability

The age-matching algorithm can be accessed via this link [https://github.com/chrjuergens/cohortagematcher]

## Author contributions

WMJW performed the PET-blot and RT-QuIC experiments, analyzed the data and wrote the manuscript. DM and LN did clinical examinations and edited the final version of the manuscript. JS designed the study and performed clinical examinations. PL and LP acquired the biopsies. FR analyzed DaT-SPECT-data. KR performed immunohistochemical experiments and analyzed results. AL performed RT-QuIC experiments. AW acquired autopsy samples. KF was involved in the study design and edited the final version of the manuscript. WJSS designed the study, acquired autopsy specimen, did the sample preparations of the biopsies, analyzed data, and edited the manuscript. All authors have approved the final version of the article.

## Acknowledgments

We thank Tatjana Pfander and Dalia Krauceviciene for excellent technical assistance. We thank Dr. Christian Jürgens for his support. We thank Dr. Manuel Buttini, who was supported by a donation from the Jean Think Foundation, Luxembourg, for his support. Prothena kindly provided the monoclonal antibody 5C12.

We mourn the unexpected death of Prof. Jörg Spiegel, who initiated this study.

This study was funded by in-house support of the Saarland University. The funder played no role in study design, data collection, analysis and interpretation of data, or writing this manuscript.

## References

1. Tysnes OB, Storstein A. Epidemiology of Parkinson’s disease. J Neural Transm (Vienna) 2017;124:901–905.

2. Lang AE. In pursuit of prodromal Parkinson’s disease. Lancet Neurol 2015;14:27–28.

3. Breen DP, Evans JR, Farrell K, Brayne C, Barker RA. Determinants of delayed diagnosis in Parkinson’s disease. J Neurol 2013;260:1978–1981.

4. Saunders-Pullman R, Wang C, Stanley K, Bressman SB. Diagnosis and referral delay in women with Parkinson’s disease. Gend Med 2011;8:209–217.

5. Cervantes Arriaga A, Rodríguez Violante M, Camacho Ordóñez A, et al. Time from motor symptoms onset to diagnosis of Parkinson’s disease in Mexico. Gaceta Médica de México 2014;150.

6. Cervantes-Arriaga A, Sarabia-Tapia C, Esquivel-Zapata O, et al. Pitfalls and caveats in the diagnostic pathway of people with ParkinsonÓ?s disease. Revista Mexicana de Neurociencia 2022;23.

7. Lay-Son L, Eloiza C, Trujillo-Godoy O. Delay in the diagnosis of Parkinson’s disease in a Chilean public hospital. 2015;143:870–873.

8. Hughes AJ, Daniel Se Fau - Kilford L, Kilford L Fau - Lees AJ, Lees AJ. Accuracy of clinical diagnosis of idiopathic Parkinson’s disease: a clinico-pathological study of 100 cases. Journal of neurology, neurosurgery, and psychiatry 1992;55 (3):181–184.

9. Beach TG, Adler CH. Importance of low diagnostic Accuracy for early Parkinson’s disease. Mov Disord 2018;33:1551–1554.

10. Kramer ML, Schulz-Schaeffer WJ. Presynaptic alpha-synuclein aggregates, not Lewy bodies, cause neurodegeneration in dementia with Lewy bodies. J Neurosci 2007;27:1405–1410.

11. Schulz-Schaeffer WJ. The synaptic pathology of alpha-synuclein aggregation in dementia with Lewy bodies, Parkinson’s disease and Parkinson’s disease dementia. Acta Neuropathol 2010;120:131–143.

12. Schneider SA, Alcalay RN. Neuropathology of genetic synucleinopathies with parkinsonism: Review of the literature. Mov Disord 2017;32:1504–1523.

13. Beach TG, Adler CH, Sue LI, et al. Multi-organ distribution of phosphorylated alpha-synuclein histopathology in subjects with Lewy body disorders. Acta Neuropathol 2010;119:689–702.

14. Concha-Marambio L, Pritzkow S, Shahnawaz M, Farris CM, Soto C. Seed amplification assay for the detection of pathologic alpha-synuclein aggregates in cerebrospinal fluid. Nat Protoc 2023;18:1179–1196.

15. Fairfoul G, McGuire LI, Pal S, et al. Alpha-synuclein RT-QuIC in the CSF of patients with alpha-synucleinopathies. Ann Clin Transl Neurol 2016;3:812–818.

16. Groveman BR, Orru CD, Hughson AG, et al. Rapid and ultra-sensitive quantitation of disease-associated alpha-synuclein seeds in brain and cerebrospinal fluid by alphaSyn RT-QuIC. Acta Neuropathol Commun 2018;6:7.

17. Okuzumi A, Hatano T, Matsumoto G, et al. Propagative alpha-synuclein seeds as serum biomarkers for synucleinopathies. Nat Med 2023;29:1448–1455.

18. Donadio V, Wang Z, Incensi A, et al. In Vivo Diagnosis of Synucleinopathies: A Comparative Study of Skin Biopsy and RT-QuIC. Neurology 2021;96:e2513–e2524.

19. Vascellari S, Orru CD, Caughey B. Real-Time Quaking-Induced Conversion Assays for Prion Diseases, Synucleinopathies, and Tauopathies. Front Aging Neurosci 2022;14:853050.

20. Adler CH, Dugger BN, Hentz JG, et al. Peripheral Synucleinopathy in Early Parkinson’s Disease: Submandibular Gland Needle Biopsy Findings. Mov Disord 2016;31:250–256.

21. Chahine LM, Beach TG, Brumm MC, et al. In vivo distribution of alpha-synuclein in multiple tissues and biofluids in Parkinson disease. Neurology 2020;95:e1267–e1284.

22. Adler CH, Dugger BN, Serrano G, et al. Submandibular gland biopsy for the diagnosis of Parkinson disease. Neuropathol Exp Neurol 2013;72(2):130–136.

23. Manne S, Kondru N, Jin H, et al. alpha-Synuclein real-time quaking-induced conversion in the submandibular glands of Parkinson’s disease patients. Mov Disord 2020;35:268–278.

24. Höglinger GU, Adler CH, Berg D, et al. A biological classification of Parkinson’s disease: the SynNeurGe research diagnostic criteria. Lancet Neurol 2024;23:191–204.

25. Postuma RB, Berg D, Stern M, et al. MDS clinical diagnostic criteria for Parkinson’s disease. Mov Disord 2015;30:1591–1601.

26. Bargar C, Wang W, Gunzler SA, et al. Streamlined alpha-synuclein RT-QuIC assay for various biospecimens in Parkinson’s disease and dementia with Lewy bodies. Acta Neuropathol Commun 2021;9:62.

27. Pinder P, Thomzig A, Schulz-Schaeffer WJ, Beekes M. Alpha-synuclein seeds of Parkinson’s disease show high prion-exceeding resistance to steam sterilization. J Hosp Infect 2021;108:25–32.

28. Haqparwar J, Pepe A, Fassbender K, et al. Reduced MIBG accumulation of the parotid and submandibular glands in idiopathic Parkinson’s disease. Parkinsonism Relat Disord 2017;34:26–30.

29. Rizzo G, Copetti M, Arcuti S, Martino D, Fontana A, Logroscino G. Accuracy of clinical diagnosis of Parkinson disease: A systematic review and meta-analysis. Neurology 2016;86 (6):556–576.

30. Borghammer P, Just MK, Horsager J, et al. A postmortem study suggests a revision of the dual-hit hypothesis of Parkinson’s disease. NPJ Parkinsons Dis 2022;8:166.

31. Wang Z, Becker K, Donadio V, et al. Skin alpha-Synuclein Aggregation Seeding Activity as a Novel Biomarker for Parkinson Disease. JAMA Neurol 2020;78:1–11.

32. Shahnawaz M, Mukherjee A, Pritzkow S, et al. Discriminating alpha-synuclein strains in Parkinson’s disease and multiple system atrophy. Nature 2020;578:273–277.

33. Shahnawaz M, Tokuda T, Waragai M, et al. Development of a Biochemical Diagnosis of Parkinson Disease by Detection of alpha-Synuclein Misfolded Aggregates in Cerebrospinal Fluid. JAMA Neurol 2017;74:163–172.

34. Simuni T, Chahine LM, Poston K, et al. A biological definition of neuronal alpha-synuclein disease: towards an integrated staging system for research. Lancet Neurol 2024;23:178–190.

