## supplemental table 1 for "Evaluation of submandibular gland biopsies with RT-QuIC in Parkinson’s disease under investigation"

***Supplemental table 1****: Diagnosis and availability of frozen and paraffin-embedded tissue of autopsy cases.*

| **Autopsy No.** | **Diagnosis** | **FFPE- SMG** | **frozen SMG** | **frozen brain** |
| --- | --- | --- | --- | --- |
| #A1 | PD | ✓ | ✓ | ✓ |
| #A2 | DLB | ✓ | ✓ | ✓ |
| #A3 | DLB | ✓ | ✓ | ✓ |
| #A4 | PD | ✓ | ✓ | ✓ |
| #A5 | PD | ✓ | ✓ |  |
| #A6 | PSP | ✓ | ✓ | ✓ |
| #A7 | ALS | ✓ | ✓ |  |
| #A8 | lymphocytic encephalitis | ✓ | ✓ | ✓ |
| #A9 | AD |  | ✓ | ✓ |
| #A10 | PSP | ✓ | ✓ |  |
| #A11 | MID |  | ✓ |  |
| #A12 | PD |  | ✓ | ✓ |

FFPE = formalin-fixed paraffin-embedded, SMG = submandibular gland, PD = Parkinson´s disease, DLB = dementia with Lewy bodies, ALS = amyotrophic lateral sclerosis, AD = Alzheimer´s disease, PSP = Progressive supranuclear palsy, MID = multiple infarct dementia
